# Two doses of the mRNA BNT162b2 vaccine reduce severe outcomes, viral load and secondary attack rate: evidence from a SARS-CoV-2 Alpha outbreak in a nursing home in Germany, January-March 2021

**DOI:** 10.1101/2021.09.13.21262519

**Authors:** Emily Dorothee Meyer, Mirco Sandfort, Jennifer Bender, Dorothea Matysiak-Klose, Achim Dörre, Gerhard Bojara, Konrad Beyrer, Wiebke Hellenbrand

## Abstract

A SARS-CoV-2 Alpha outbreak was detected in a nursing home after residents and staff had completed vaccination with BNT162b. In a retrospective cohort study, we estimated an age-adjusted vaccine effectiveness of 88% [95% confidence interval (95%CI) 41-98%] against hospitalization/death. Ct values at diagnosis were higher with longer intervals since the second vaccination [>21 vs. ≤21 days: 4.82 cycles, 95%CI: 0.06-9.58]. Secondary attack rates were 67% lower in households of vaccinated [2/9 (22.2%)] than unvaccinated infected staff [12/18 (66.7%); p=0.046]. Vaccination reduced the risk of severe outcomes, Ct values and transmission, but not fully. Non-pharmaceutical interventions remain important for vaccinated individuals.

## Background

While COVID-19 case-fatality was <0.1% in under-50-year-olds [1], it was 13% in outbreaks of SARS-CoV-2 in nursing homes from January 2020 to February 2021 in Germany [2]. Hence, nursing homes were prioritized for COVID-19 vaccination which began in Germany in December 2020 [3]. We report on a SARS-CoV-2 Alpha (B.1.1.7) outbreak among residents and staff of a nursing home in Germany, of whom some were vaccinated with two doses of BNT162b. This study describes the epidemiology of the outbreak, the undertaken control measures and the vaccine effectiveness (VE) against SARS-CoV-2 Alpha infections, disease and severe outcomes (hospitalization or death) and the vaccine effects on viral load and secondary transmission.

## Study design

Cases were defined as residents (either permanent or day-care) or staff who had a positive SARS-CoV-2-PCR between early January 2021 (symptom onset of first case denoted as day 0) and mid-March 2021 (2 weeks after diagnosis of the last case, day 74). In a retrospective cohort study, we included all residents and staff who attended the nursing home during the same time period. We compared attack rates (AR) with Chi-squared or Fisher’s exact tests. We estimated vaccine effectiveness (VE) as VE=1-RR, where RR denotes the relative risk for the respective outcome in vaccinated vs. unvaccinated individuals, calculated by Poisson regression. Considered outcomes were SARS-CoV-2 infection, symptomatic infection and severe courses (defined as hospitalization or death from COVID-19). Using linear regression, we analyzed the effect of the time interval between the second vaccine dose and viral load at diagnosis (using Ct value for the ORF1AB gene at the first positive PCR as a proxy). Unvaccinated cases were assigned an interval of 0 days. We assessed secondary attack rates (SAR) among household members of SARS-CoV-2-positive staff, who were tested twice during their quarantine. One household outside the administrative district was excluded because data was unavailable. Secondary cases were defined as SARS-CoV-2 PCR-positive household members diagnosed 1-14 days after the diagnosis of the corresponding staff index case.

## Ethical statement

This outbreak investigation was conducted in accordance to paragraph 4 of the German Protection against Infection Act. Therefore, this investigation was exempt from additional institutional review.

## Study setting

The nursing home comprised one day-care and seven permanent care wards with 128 members of staff, 100 residents in permanent care and 24 persons in day-care. Ninety-five/124 (77%) residents and 72/128 (56%) staff members were vaccinated with BNT162b in early and late January 2021, with an inter-dose interval of three weeks. Median age was 49 years among staff (inter-quartile range (Q25-Q75): 32-58) and 87 years among residents (Q25-Q75: 83-92). Among residents, 97/124 (77%) and among staff, 113/128 (88%) were female.

### Measures in place before the outbreak

All staff had to conduct daily rapid antigen detection tests (RADT). Residents were tested related to incidences, e.g. when becoming symptomatic. All visitors of the nursing homes could only enter with a negative RADT of the same day. Staff and visitors had to wear FFP2 standard masks inside the nursing home. Staff was assigned in teams to designated wards and rotation between wards was minimized. However, this was not possible for night shifts and social workers.

## The epidemiology of the outbreak

After detecting the first SARS-CoV-2 infection in a permanent care resident in early February 2021 (day 30) and previous, sporadic cases among four staff since early January 2021 (day 0), an outbreak investigation was initiated. Between early January 2021 (day 0) and 14 days after the detection of the last case in mid-March 2021 (day 74), 50 SARS-CoV-2 cases were detected, of which 35 were symptomatic (70%; 16/35 (46%) vaccinated). Four residents (1/4 (25%) vaccinated) were hospitalized and five died (2/5 (40%) vaccinated) from or with COVID-19. The crude AR among residents (AR=27%, 34/124) was higher than among staff (AR=12%, 16/128, p<0.01). Typing of PCR samples for variants of concern detected variant Alpha in 27/28 samples.

### The course of the outbreak

A kitchen staff, working in a kitchen serving all sections, developed symptoms on day 0, worked for two more days prior to isolation with no reported contacts to other care sections. Between day 8 and day 23, mainly the day-care was affected with nine detected cases (eight residents and one member of staff) and was therefore closed. In the stationary care, two members of staff developed symptoms on day 28 and a resident was RADT-positive on day 30. Subsequently, the outbreak spread in the permanent care wards, see Figure 1 and 2.

**Figure 1:**
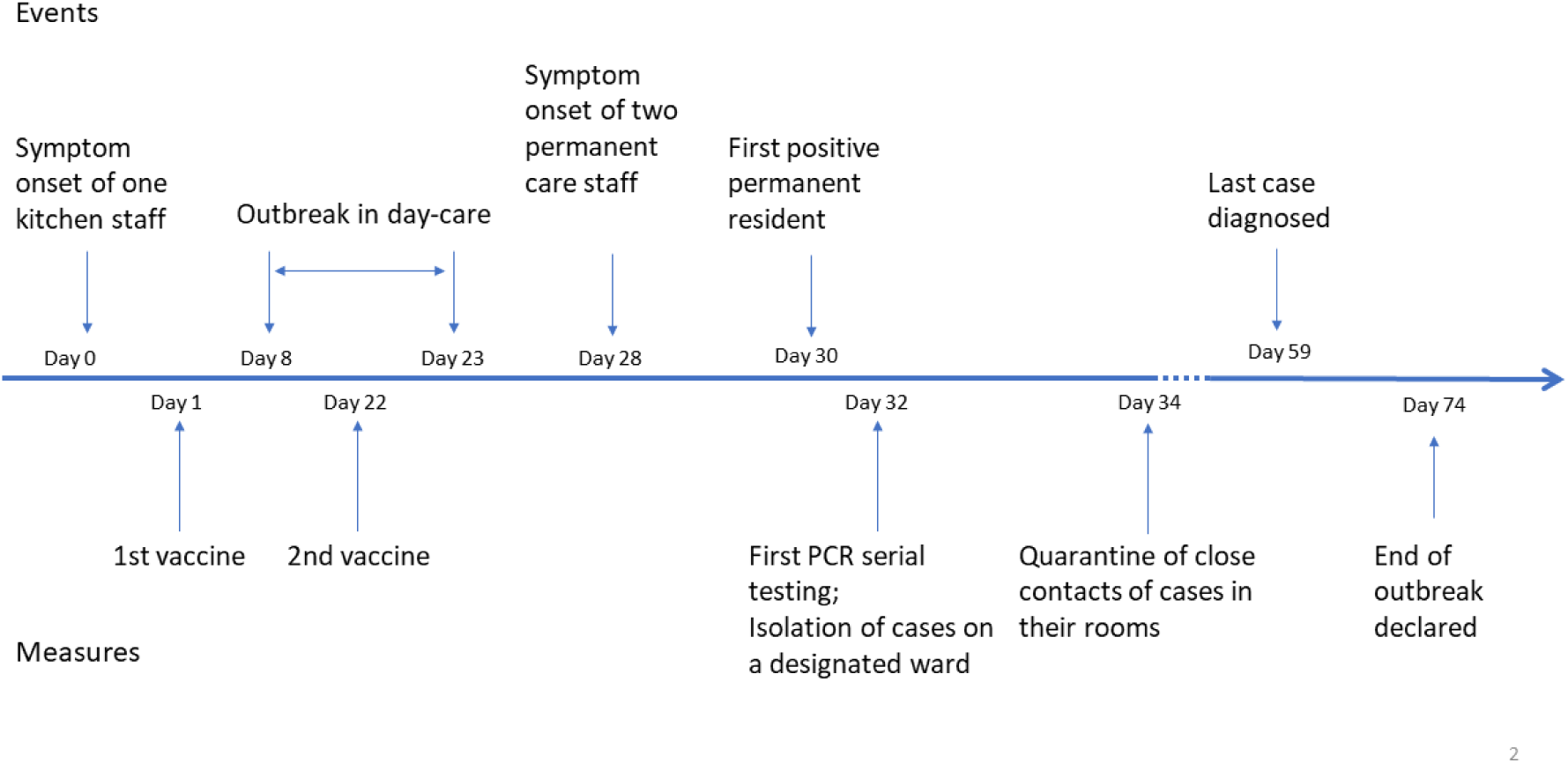
Timeline of events and measures during a SARS-CoV-2 Alpha outbreak in a nursing home in Germany, Jan-Mar 2021.

**Figure 2:**
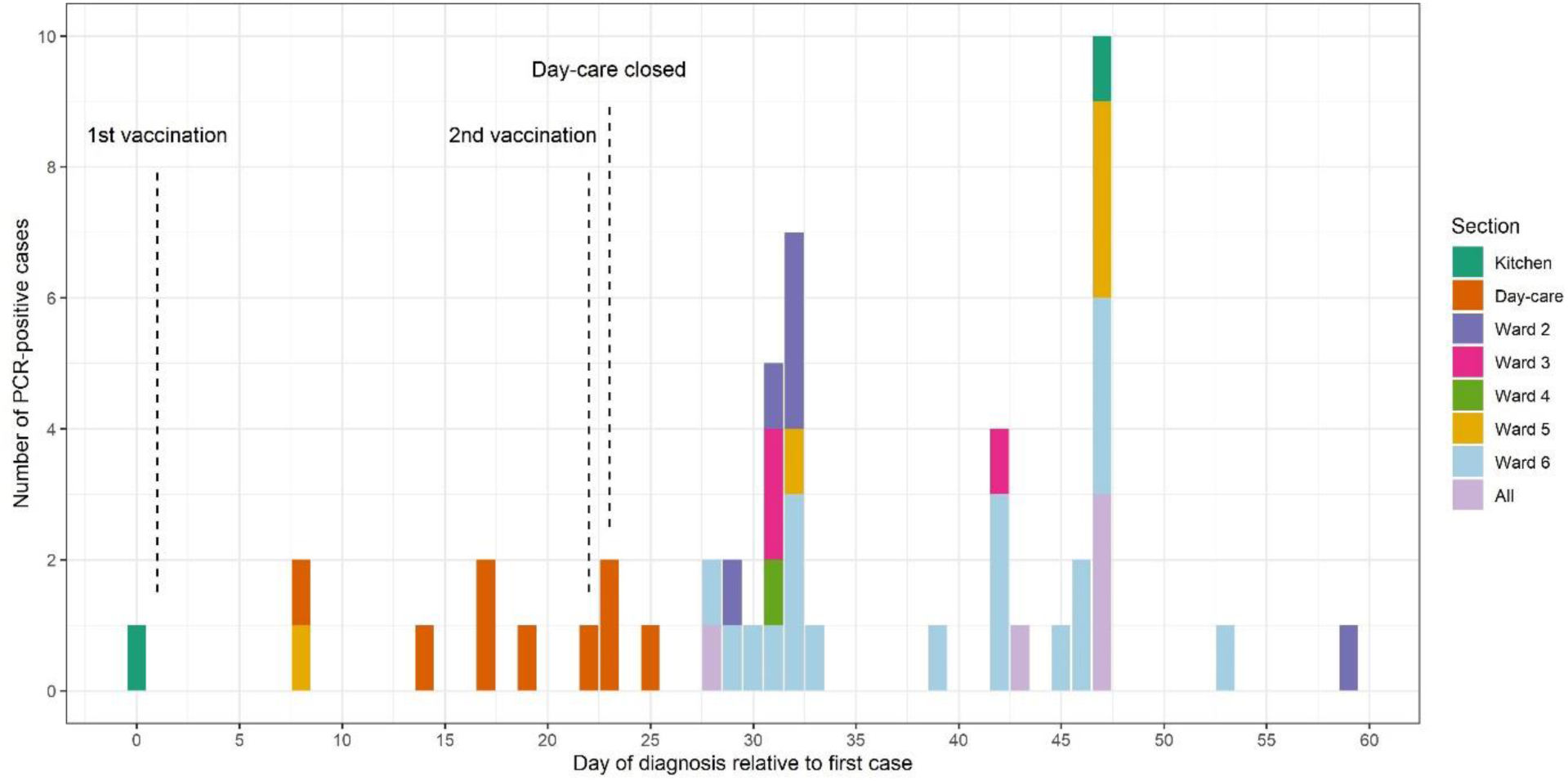
Cases of SARS-CoV-2 Alpha by time of diagnosis during an outbreak in a nursing home in Germany, Jan-Mar 2021. Color represents the area of residence or activity within the nursing home (“all” if no designation to a specific ward). Date of diagnosis defined as the earlier date of either symptom-onset or sampling of a positive test.

### Potential sources of the outbreak

All members of the vaccination team were tested negative with daily RADT and with weekly PCR. Therefore, it seems unlikely that the vaccination team was the source of the outbreak. Initially, no links between the day-care and the stationary care were reported. However, the contact tracing information revealed that an external health-care worker (ID-5160, see Figure 3) visited a highly infectious case (ID-2640, Ct value 11) from the day-care on day 10 and a person from the permanent care (ID-870) on day 12 (who tested positive later), suggesting that they visited ID-870 within the infectious period. ID-5160 was tested positive with an RADT on day 13. Potentially, the ID-5160 represents a link between these two sections. However, no sequencing data was available to further delineate possible transmission chains. The other 165 visitors of the nursing home did not have timely relevant SARS-CoV-2 infections and were therefore excluded as potential sources of infections.

**Figure 3:**
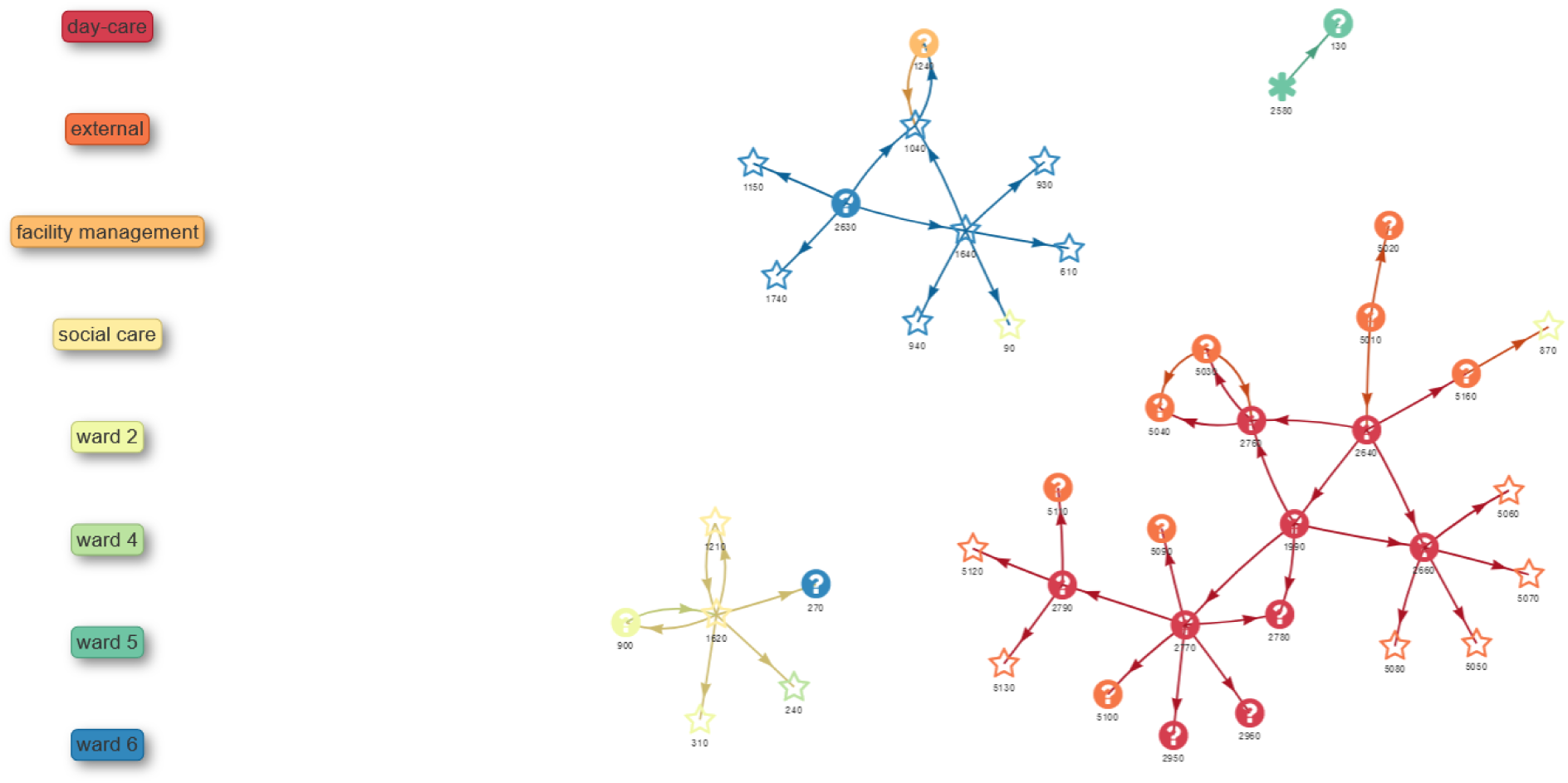
Contact network of PCR-positive cases of a SARS-CoV-2 Alpha outbreak in a nursing home in Germany, Jan-Mar 2021. Star: PCR typing: Alpha Asterix: PCR typing: wild type Question mark: No typing performed Number below symbol: ID Color represents the area of residence or activity within the nursing home or external visitors or household members

**Figure 4:**
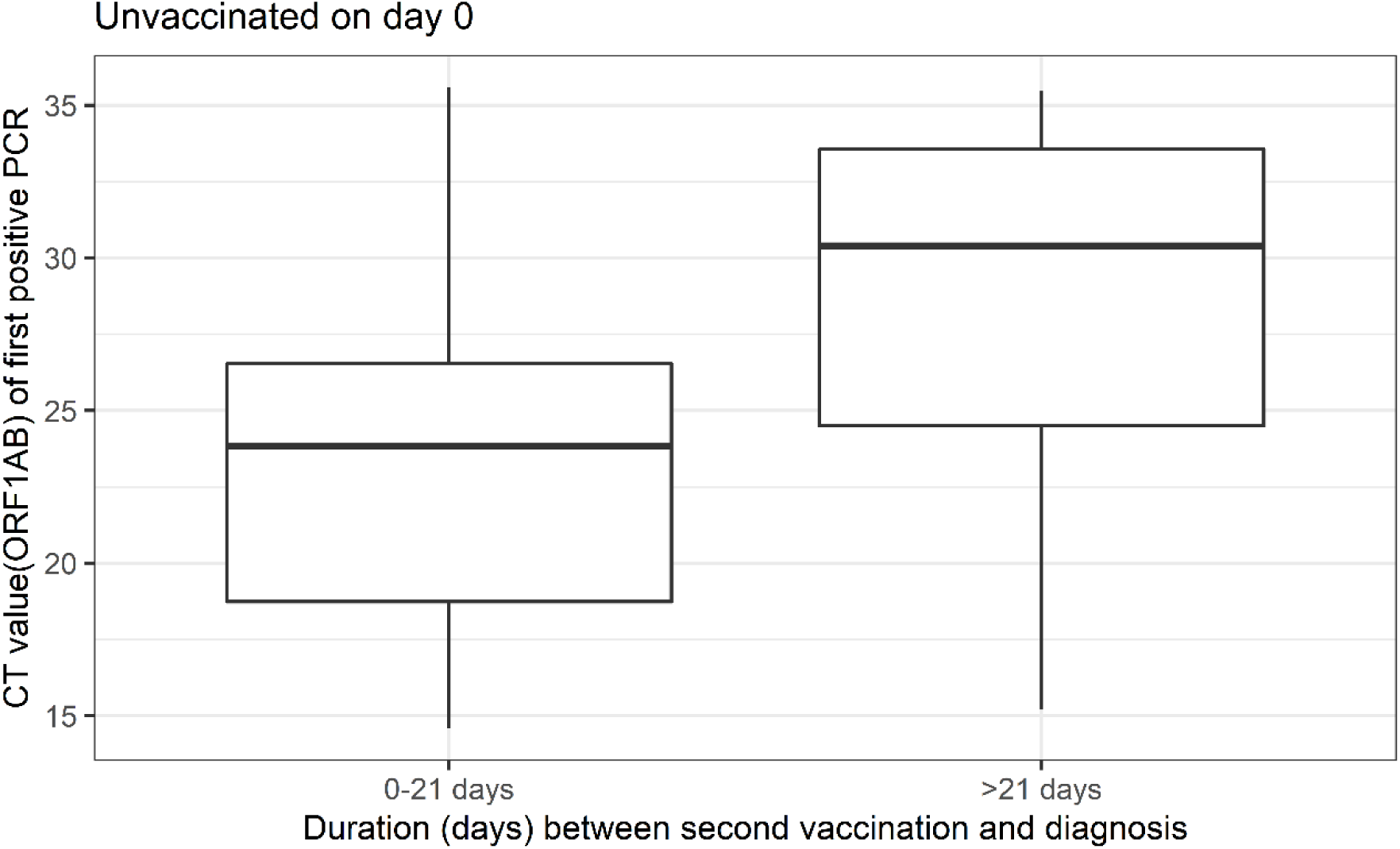
Ct-Value (ORF1AB gene) of the first positive PCR by interval between second vaccination and diagnosis during a SARS-CoV-2 Alpha outbreak in a nursing home in Germany, Jan-Mar 2021.

### Outbreak management

On day 32, 4 days after the first member of staff of the stationary care was symptomatic, regular PCR serial testing was implemented for all residents and staff every 5 days until two consecutive PCR serial tests had negative results only. All symptomatic or PCR-positive residents were isolated as a cohort on a designated ward. Non-cases could move within their ward but contacts between wards were minimized. Close contacts were quarantined in their rooms. Case isolation ended once cases were asymptomatic and were PCR-negative at the earliest after 14 days. No visitors were allowed between day 31 and day 74.

## Vaccine effectiveness

Among 29 vaccinated cases, the date of diagnosis (defined as the earlier date of either symptom-onset or sampling of a positive test) was 7-11 days after the second vaccine in 14 cases (48%, all residents), while 15 (52%) were diagnosed 20 or more days post vaccination. SARS-CoV-2 infections were diagnosed less frequently among vaccinated than unvaccinated residents (Table 1, p=0.46) and staff (Table 1, p=0.06). Age-adjusted VE ≥7 days after two doses of BNT162b was 45% (0-69%, p=0.048) against infection. Among residents and staff, vaccinated cases were less symptomatic than unvaccinated cases (Table 1, residents: p=0.04, staff: p<0.01). Age-adjusted VE was 68% (36-84%; p<0.01) against disease. Of 50 cases, four were hospitalized (1/4 (25%) vaccinated) and five (2/5 (40%) vaccinated) died (all residents). Age-adjusted VE ≥7 days after completed vaccination was 88% (37-98%; p=0.01) against severe outcomes.

**Table 1:** Outcomes of a SARS-CoV-2 Alpha outbreak in a nursing home in Germany, Jan-Mar 2021, stratified by vaccination status among residents, staff and all.

|  | Vaccination |  |  | Case |  | Symptomatic |  | Hospitalization |  | Death |  |
| --- | --- | --- | --- | --- | --- | --- | --- | --- | --- | --- | --- |
|  |  | n | % | n | AR | n | % of cases | n | % of cases | n | % of cases |
| Residents | No | 29 |  | 10 | 34% | 8 | 80% | 3 | 30% | 3 | 30% |
|  | Yes | 95 | 77% | 24 | 25% | 13 | 54% | 1 | 4% | 2 | 8% |
| Staff | No | 56 |  | 11 | 20% | 11 | 100% | 0 | 0% | 0 | 0% |
|  | Yes | 72 | 56% | 5 | 7% | 3 | 60% | 0 | 0% | 0 | 0% |
| Total | No | 85 |  | 21 | 25% | 19 | 90% | 3 | 14% | 3 | 14% |
|  | Yes | 167 | 66% | 29 | 17% | 16 | 55% | 1 | 3% | 2 | 7% |

Attack rate (AR) with the strata size as denominator. Symptomatic cases, hospitalization and deaths with respective number of cases as denominator.

Age confounded the association between vaccination status and infection, disease and severe outcomes, changing effect estimates by 21% (from 0.70 to 0.55), 20% (from 0.40 to 0.32) and 50% (from 0.24 to 0.12), respectively (Table 2). Gender was not associated with risks for infection (p=0.64) or disease (p=0.69), but women exhibited a lower risk for severe outcomes (p=0.07, Table 2), however not after adjusting for age and vaccination status [RR=0.34, 95%CI: 0.06-1.85, p=0.20].

**Table 2.**
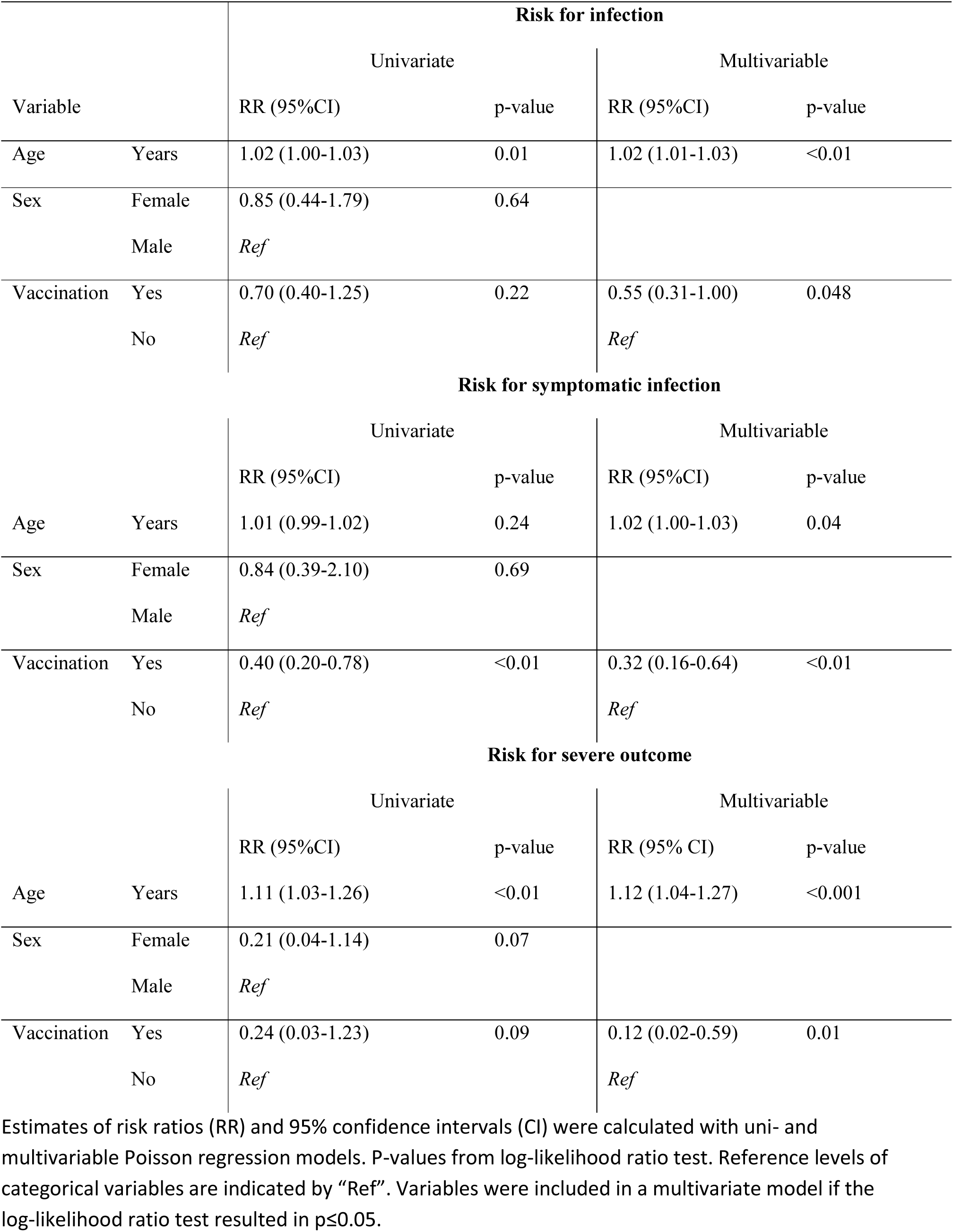
Risk for SARS-CoV-2 infection, risk for symptomatic infection and risk for severe outcome (hospitalization or death) during a SARS-CoV-2 Alpha outbreak in a nursing home in Germany, Jan-Mar 2021.

Among symptomatic cases, no association of the time interval between the second vaccine dose and diagnosis with the risk for severe outcomes was observed (time in days: IRR=0.93, 95%CI: 0.76 – 1.06, p=0.34; 7-14 days vs. >14 days: IRR=0.78, 95%CI:0.10-15.81, p=0.83), adjusted for age.

## Vaccination reduced the viral load

Ct values at diagnosis were on average 3.04 cycles (Table 3, p=0.28) higher among vaccinated than unvaccinated cases. The Ct value increased with time since the second vaccine dose (Table 3 and Figure 3). Age (p=0.79) and sex (p=0.61) was not associated with the Ct value (Table 3).

**Table 3:** Associations with the Ct value of the PCR at diagnosis in a SARS-CoV-2 Alpha outbreak in a nursing home in Germany, Jan-Mar 2021.

| Variable |  | Coefficient | 95%CI | p-value |
| --- | --- | --- | --- | --- |
| Age | Δ years | -0.02 | -0.15 – 0.12 | 0.80 |
| Sex | Female | 1.53 | -4.81 – 7.87 | 0.63 |
|  | Male | Ref |  |  |
| Symptoms | No | 6.32 | 1.89 – 10.75 | **<0.01** |
|  | Yes | Ref |  |  |
| Vaccination | Yes | 3.04 | -2.86 – 8.93 | 0.30 |
|  | No | Ref |  |  |
| Time interval between second vaccine and diagnosis | Δ days | 0.23 | 0.01 – 0.45 | **0.04** |
|  | > 14 days | 3.53 | -1.15 – 8.22 | **0.13** |
|  | ≤ 14 days | Ref |  |  |
|  | > 21 days | 4.82 | 0.06 – 9.58 | **0.047** |
|  | ≤ 21 days | Ref |  |  |

The results were calculated with univariate linear regression models. Δ: indicates a numerical variable. 95%CI: 95% confidence interval; P-values are based on Wald tests.

## Vaccination reduced secondary transmission in households

We analyzed 14 households of SARS-CoV-2-positive staff (five vaccinated, nine unvaccinated). We found two secondary cases in 1/5 (20%) households of vaccinated staff (index staff case was diagnosed 25 days after the second vaccination) and 12 secondary cases in 5/9 (56%) households of infected, unvaccinated staff. For calculating the adjusted SAR, we excluded household members with a PCR-confirmed SARS-CoV-2 infection <6 months prior or who quarantined separately from infected staff. Household members with a vaccinated index case had a lower SAR [2/9 (22%)] than household members of unvaccinated SARS-CoV-2-positive staff (p=0.046, Fisher’s exact test, Table 4).

**Table 4:** Secondary SARS-CoV-2 cases and secondary attack rate (SAR) in households of SARS-CoV2-positive staff, stratified by vaccination status, during a SARS-CoV-2 Alpha outbreak in a nursing home in Germany, Jan-Mar 2021.

| Vaccination<br>status of index<br>staff case | Household members |  |  |  |  | Crude Analysis |  | Adjusted Analysis |  |
| --- | --- | --- | --- | --- | --- | --- | --- | --- | --- |
|  | Total | Infected | Prior | Separate | Total | n/cN | SAR | n/adj.N | SAR |
|  | (cN) | (n) | infection | isolation | (adj.N) |  |  |  |  |
| Vaccinated | 9 | 2 | 0 | 0 | 9 | 2/9 | 22% | 2/9 | 22% |
| Unvaccinated | 22 | 12 | 2 | 2 | 18 | 12/22 | 55% | 12/18 | 67% |
| Total | 31 | 14 | 2 | 2 | 27 | 14/31 | 45% | 14/27 | 52% |

cN: crude N, adj.N adjusted N; Adjusted SAR excludes household members infected within 6 months prior to the infection of the staff index case and excludes household members who isolated separately from the staff index case.

## Discussion

In our study, the age-adjusted VE of two doses of BNT162b was moderate against infection and disease and high against severe COVID-19. Our VE estimates are lower than in a population-based cohort study conducted in Israel [4] that reported a VE of 96% against hospitalization and 93% against death. This can be explained by a higher potential for repeated contacts with infected cases and a higher median age in this outbreak setting. Furthermore, half of the vaccinated cases were diagnosed within 7-11 days after the second vaccination; thus, infection occurred prior to attaining full immunity. However, we did not observe higher effectiveness among cases with a longer interval between the second vaccine and diagnosis, in line with findings from the UK [5]. Our analysis is limited by the small sample size and inability to control for risk factors such as underlying chronic diseases and compliance with protective measures. The individual risk of infection possibly changed over time with the implementation of non-pharmaceutical control measures, such as separating cases from non-cases, thereby potentially biasing the VE results of our study.

Our VE against infection was lower than in previous studies from similar settings [4], [6], [7]. Since this cohort was PCR-tested every 5-6 days throughout the outbreak, we believe that the risk for missing asymptomatic cases was minimal while in other studies under-ascertainment of asymptomatic infection may have occurred.

We found a significantly lower Ct value among vaccinated cases ≥21 days after the second vaccine (6 weeks after the first) than among non-vaccinated cases. One study assessing a similar study population [8] found lower viral loads already four weeks after the first vaccination. However, the authors pooled different Ct values from all available PCR-positive results, which may have impacted the inter-assay comparability [9], especially considering N-gene dropouts with the Alpha variant [10].

Our results suggest that while transmission was reduced, close contacts of vaccinated persons with break-through SARS-CoV-2 infections remained at risk for infection, as shown in previous studies for household members of healthcare workers vaccinated with BNT162b2 [11]. As no samples could be sequenced, our results are limited by the inability to show that primary and secondary cases had identical viral strains. Still, these results have strong implications for policy makers because they emphasize that adhering to non-pharmaceutical interventions is still very important for vaccinated people.

We believe that the regular PCR serial testing, isolating cases on a designated ward and quarantining close contacts of cases in their rooms contributed to the successful outbreak control. It is possible that the outbreak on the day-care and the stationary ward were linked via a visiting health-care worker. However, typing revealed that at least two different strains were present in this outbreak, suggesting two introductory events of SARS-CoV-2 at minimum.

## Conclusions

Two doses of BNT162b significantly reduced the risk for SARS-CoV-2 infections, symptomatic infections, severe outcomes, viral load and secondary transmission, even within 14 days after the second dose. However, the incomplete protection emphasizes that adhering to non-pharmaceutical interventions remains important after completed vaccination. When reconsidering non-pharmaceutical measures for fully vaccinated people, policy makers need to be aware that the risk reduction for vaccination people regarding the risk for infection, severe outcomes and transmission is incomplete. Regular PCR serial testing and isolation of cases on a designated ward contributed to the outbreak control.

## Supporting information

Statement regarding additional institutional reviews

## Data Availability

Aggregated data from a limited version of the German notification system database can be retrieved via SurvStat@RKI 2.0 https://survstat.rki.de/. Detailed data are confidential and protected by German law and are available from the corresponding author upon reasonable request.

## Acknowledgement

The authors are grateful for the work and the commitment of all public health staff throughout the pandemic. The authors would like to specifically thank the Local Public Health Authority, Osnabrück, the Department of Infectious Diseases of the Public Health Agency of Lower Saxony, Hannover, Germany and the staff of the affected nursing home for their cooperation in the investigation. Emily Meyer, Mirco Sandfort and Jennifer Bender would like to thank their frontline coordinators Sybille Rehmet, Katharina Alpers and Loredana Ingrosso for their continuous support.

## Disclaimers

No conflict of interest reported. JB is a fellow of the ECDC Fellowship Programme. The views and opinions expressed herein do not state or reflect those of the ECDC. The ECDC is not responsible for the data and information collection and analysis and cannot be held liable for conclusions or opinions drawn.

