## Supplementary material for "Two doses of the mRNA BNT162b2 vaccine reduce severe outcomes, viral load and secondary attack rate: evidence from a SARS-CoV-2 Alpha outbreak in a nursing home in Germany, January-March 2021": Statement regarding additional institutional reviews

ROBERT KOCH INSTITUT

Robert Koch Institute | Seestrasse 10 | 13353 Berlin

Department for Infectious Disease Epidemiology

Dr. Osamah Hamouda

To whom it may concern

26.08.2021

**Two doses of the mRNA BNT162b2 vaccine reduce severe outcomes, viral load and secondary attack rate: evidence from a SARS-CoV-2 Alpha outbreak in a nursing home, Osnabrück, Germany, January-March 2021**

As head of the Department for Infectious Disease Epidemiology at the Robert Koch Institute (RKI), the national Public Health Institute of Germany, I hereby certify that:

The outbreak investigation presented by Meyer et al. in "**Two doses of the mRNA BNT162b2 vaccine reduce severe outcomes, viral load and secondary attack rate: evidence from a SARS-CoV-2 Alpha outbreak in a nursing home, Osnabrück, Germany, January-March 2021**" was conducted as part of the official tasks of the local public health authorities of the respective district, supported by the RKI upon official request in accordance to §4 of the German Protection against Infection Act. Therefore, this investigation was exempt from additional institutional review.

Sincerely,

Dr. Osamah Hamouda

Head of Department for Infectious Disease Epidemiology

Robert Koch Institute

Robert Koch Institute

www.rki.de

Reporting/
Processing by: M. Meyer

Extension: -5215

Address:
Seestrasse 10
13353 Berlin
Germany

The Robert Koch Institute is a federal institute within the portfolio of the Federal Ministry of Health
